# Variation in price of cardiovascular and diabetes medicine in Indonesia, and relationship with quality: a mixed methods study in East Java

**DOI:** 10.1101/2022.11.24.22282722

**Authors:** Elizabeth Pisani, Aksari Dewi, Anna Palagyi, Devarsety Praveen, Bachtiar Rifai Pratita Ihsan, Ayuk Lawuningtyas Hariadini, Diana Lyrawati, Sujarwoto, Asri Maharani, Gindo Tampubolon, Anushka Patel

## Abstract

Lower-middle income Indonesia, the world’s fourth most populous country, has struggled to contain costs in its mandatory, single-payer public health insurance system since the system’s inception in 2014. Public procurement policies radically reduced prices of most medicines in public facilities and the wider market. However, professional associations and the press have questioned the quality of these low-cost, unbranded generic medicines.

We collected 204 samples of 4 cardiovascular and 1 antidiabetic medicine from health facilities and retail outlets in East Java. We collected amlodipine, captopril, furosemide, simvastatin and glibenclamide, sampling to reflect patients’ likelihood of exposure to specific brands and outlets. We recorded sales prices and maximum retail prices, and tested medicines for dissolution and percent of labelled content, using high-performance liquid chromatography. We conducted in-depth interviews with supply chain actors.

All samples, including those provided free in public facilities, met quality specifications.

Most manufacturers make both branded and unbranded medicines. Retail prices varied widely. The median ratio of price to the lowest price for an equivalent product was 5.1, and a few brands sold for over 100 times the minimum price. Prices also varied between outlets for identical products, as retail pharmacies set prices to maximize profit. Since very low-cost medicines were universally available and of good quality, we believe richer patients who chose to buy branded products effectively protected medicine quality for poorer patients in Indonesia, because manufacturers cross-subsidize between branded and unbranded versions of the same medicine.

## Background

As middle-income countries expand their efforts to provide Universal Health Coverage, pressure on government health budgets has increased.^1^ Cost-containment measures often include procurement policies that seek to bring down the price of medicines in the public system, including by increasing the proportion of unbranded generic medicines used.^2^ However, sharp falls in prices have, in some cases, led to questions being raised about the quality of publicly procured medicines.^3^

Previous studies indicate that physicians and patients continue to question the quality of unbranded generic and other low-cost medicines, despite substantial and growing evidence that they are therapeutically equivalent or superior to originator medicines and other branded products.^4–6^ Global reviews show that this is especially likely to be the case in low- and middle-income countries, where medicines provided for free in public health systems are particularly mistrusted.^7–9^

In Indonesia, the world’s fourth most populous nation, a national medicines procurement system was introduced in 2014 in support of a new, mandatory, single-payer health insurance system that, by mid-2022, covered over 80% of the population.^10^ The single-winner auction system, known as e-catalog, created intense competition among domestic producers of unbranded generic medicines, and drove prices for common medicines for chronic diseases sharply lower. Nearly 80% of the medicine procured in 2017 through e-catalog had fallen in price compared to 2013; prices of 39% of these medicines fell by more than 50%.^11^ The downward trend has continued in more recent years. For example, the price of blood pressure-lowering medicine amlodipine fell from 440 rupiah per 10mg tablet in 2013 to 70 rupiah by 2022 (US$ 0.045 to 0.005), while cholesterol control medicine simvastatin, fell from 180 rupiah per 10mg tablet to 68 rupiah (US$ 0.018 to 0.005) over the same period.^12^ Falling prices have led one multinational generics producer to withdraw from the Indonesian market entirely.^13^ The domestic industry association has warned that unsustainably low prices may threaten the quality and sustainability of supply, and in the early years of the new procurement system, newspapers and consumer associations regularly called into question the quality of medicines provided in public clinics and hospitals.^14–17^

Prices for many unbranded generics (also referred to as International Non-proprietary Name or INN generics) in the private market fell in tandem with public procurement prices. However, many domestic pharmaceutical companies that hold market authorisations to sell these unbranded products also sell branded versions of the same medicines. While under Indonesian regulations, these should be formulated identically with the unbranded product registered to the same market authorisation holder, they sell at many times the price.

Patients who are not insured, or who do not want to queue at public facilities for free medicines, can pay for medicines at retail pharmacies. Patients may buy more expensive medicines because they are prescribed by a doctor or suggested by a health care worker or pharmacist, who may be rewarded with increased profits or through pharmaceutical company incentives if the patient takes a more expensive medicine.^18^ They may also choose more expensive medicines, even when a cheaper version is offered, because they associate higher prices with better quality.^13^

The maximum retail price is printed on the primary packaging of all medicines, in accordance with Indonesian regulations. Pharmacuetical companies are free to set the maximum retail price for branded medicines for which they hold market authorisations at any price of their choosing. Regulations passed in 2015 cap the maximum retail price for unbranded generics at the public procurement price plus 28%.^19^ In practice, prices charged to patients are not always in line with the maximum retail price; products are sold both at above and below that price.

There has, to our knowledge, been only limited description of price variation in the Indonesian medicine market,^20^ and no independent investigation of the association between medicine price and medicine quality in Indonesia. In this study, we investigate the relationship between price and quality for 204 samples of 5 medicines for cardiovascular disease (CVD) and diabetes in Malang district, East Java, Indonesia. We further describe the variation in medicine prices by branded status, brand identity and point dispensing to patients.

## Methods

The study centred on 8 villages in Malang district, a semi-rural district in Indonesia’s second most populous province, which were selected because they were the site of a household census of the prevalence of risk for cardiovascular disease.^21^ The study methods are reported in detail elsewhere, according to MEDQUARG guidelines.^22^ The MEDQUARG checklist is available in the study archive.^23^

Briefly, we designed an exposure-based sample frame. We selected the 5 medicines that patients at high risk for cardiovascular disease most commonly reported consuming in an earlier household census. ^21,24^ These were cardiovascular medicines amlodipine, captopril, furosemide and simvastatin, as well as the anti-diabetes medicine glibenclamide. (For brevity, we refer to these as “study medicines” throughout the paper.) We then triangulated data from the patient survey with data from pharmacies, the public procurement system and pharmaceutical marketing tracking systems, and constructed a sample frame based on the estimated likelihood that patients at high risk for cardiovascular disease and diabetes in the study area would consume a particular medicine from a particular source. Details of sample frame construction can be found at https://doi.org/10.7910/DVN/EBQYUB, File 2.

### Medicine outlets

We collected samples from the district medicine warehouse (1/1), which supplies all public primary health care clinics (known as *pusat kesehatan masyarakat* or *puskesmas*) and their outreach services; the district hospital (1/1); private doctors and midwives (8/30 of those reporting selling study medicines); private pharmacies (55/75 pharmacies) and over-the-counter medicine shops (2/3 of those found to sell study medicines). In addition, we collected any medicines provided for free to patients in *puskesmas* that were procured directly from distributors or pharmacies, using capitation funds (2/2).

We note that all of the study medicines are regulated as prescription-only. This means that private pharmacies are only allowed to sell them to patients with prescriptions. Private health care providers and over-the-counter medicine shops are not technically allowed to sell them in the study area. However, they commonly do so, and were thus included in the sample. For more details see ^22^.

### Sample collection strategy

All samples were collected between February – May 2021. We sampled overtly from public facilities, taking one sample of every available study medicine. In pharmacies and over-the-counter medicine shops, we used mystery shoppers posing as patients or family or friends of patients. At each outlet, shoppers requested a single medicine, or a combination consistent with common clinical needs. In order to approximate the market distribution of medicine price points, they signalled their desire for cheaper or more expensive medicines using phrases such as “*Minta yang terjangkau*” (I want something affordable) or *“Ada yang paten?*” (Do you have anything “patent”? -- the term commonly used in Indonesia to signify a premium product.)

If the sample frame called for clinically incompatible combinations, or repetitions (for example cheap and expensive versions of the same product) from a single outlet, different mystery shoppers were used.

### Sample handling and testing

All the study medicines are normally packaged in strips/blisters of 10 tablets. We aimed to collect 40 tablets per sample but accepted a minimum of 30 tablets.

On exiting the outlet, collectors put each sample in a sealable plastic bag marked with a pre-printed barcode. The barcode was scanned and field-related data were entered into a form pre-loaded onto the shoppers’ mobile phones, using open-source KoboCollect software.^26^ Further data entry, including product photographs and details of market authorisation holder, manufacturer, registration number and expiry date took place at the end of the day, using a second form linked by the same barcode. The ODK-format data collection forms are available at https://doi.org/10.7910/DVN/EBQYUB Files 03 and 04.

Research team members inspected packaging visually. No reference packaging was available for comparison, so visual inspection, using a magnifying glass as necessary, was limited to checking for anomalies such as mis-spellings, and discrepancies in formatting of batch numbers and expiry dates.

Samples were stored in a temperature-controlled environment for an average of 21 days, batched and sent (with a temperature logger) for testing to PT Equilab International, an ISO/IEC 17025-certified private laboratory in Jakarta, according to USP 42 NF 37 monograph and using USP reference standards. Methods were validated for all APIs before testing. The full protocols for each molecule are available at https://doi.org/10.7910/DVN/EBQYUB, Files 09-14.

Briefly: laboratory staff inspected tablets visually, noting shape, colour, lettering and other defining characteristics. Chemical analysis was performed for determination of identity, assay (% of labelled active ingredient) and dissolution (% of labelled active ingredient in the tablet dissolved over time). For all APIs, assay testing was by high-performance liquid chromatography, (HPLC -UV; Waters, Aliance 2695with UV Detector 2489 for amlodipine, glibenclamide, furosemide and simvastatin; Waters, Aliance 2695 with Photodiode Array Detector 2996 for captopril)), while dissolution was by Spectrophotometer-UV/VIS (Shimadzu UV-1800) with the exception of glibenclamide, where dissolution was tested by HPLC (Waters, Aliance 2695 with UV Detector 2489).

Assay testing was duplicated; the reported result is the average of the 2 tests.

We could not afford to test for uniformity or impurities.

Staff conducting the tests differed from those handling the packaged product, but could see any defining marks on tablets or capsules. Testing took place April – August 2021, an average of 95 days after sample collection.

Results from the certificate of analysis were entered into a database by study staff, using the sample barcode as identifier. Raw dissolution data were added to the database at a later date, delaying stage 2 dissolution. Where necessary, this was undertaken in March 2022.

Table 1 shows the definitions used for compliance with specifications, following USP 42 NF 37 limits.

**Table 1.** Limits of compliance, United States Pharmacopeia 42 [% of declared content].

| API | Assay (%) | Dissolution [Q] (%) | Stage 1 dissolution [Q+5] (%) |
| --- | --- | --- | --- |
| Amlodipine | 90-110 | 75 | 80 |
| Captopril | 90-110 | 80 | 85 |
| Furosemide | 90-110 | 80 | 85 |
| Glibenclamide | 90-110 | 70 | 75 |
| Simvastatin | 90-110 | 75 | 80 |

If any one of 6 pills included in stage 1 dissolution fell below the Stage 1 threshold of Q+5, we continued to stage 2 testing using additional 6 tablets. The sample was considered out of specification if:

- The assay fell outside the stated limits OR
- Any single tablet fell below the Q threshold -25 in dissolution testing OR
- Any 3 tablets fell below Q threshold -15 in dissolution testing OR
- The average of 12 tablets fell below the Q threshold in stage 2 dissolution testing.

### Panel pricing data

With the written consent of pharmacy owners, we collected monthly data from 2 private pharmacies on all brands of stocked study medicines between March and October 2021. One was a local branch of a national chain, the other an independent pharmacy. From their stock management systems, pharmacists provided us with volumes received and volumes dispensed by brand, as well as buying prices and selling prices.

We calculated the profits by brand by multiplying sales volumes by margin (selling price - buying price). We estimated the list price for each medicine by subtracting the maximum margin of 28% (10% tax and 18% “service fee”) allowed by Indonesian regulations ^19^ from the maximum retail price, then estimated the percent discount at which each product was acquired by comparing the buying price to the list price.

### Quantitative data analysis, medicine quality and pricing survey

The field data form, product data form and the laboratory data were merged on barcode number using Stata 17.0 software. Stata 17.0 was also used for reproducible cleaning and coding, and to generate descriptive statistics and graphs.

In order to be able to compare price variation between molecules sold at different prices, we calculated the ratio of each sample price to the lowest recorded price paid for the medicines. In most pricing analysis we excluded “zero” retail prices -- those medicines provided free through the public health system. In the case of the district hospital, which provides medicines free to insured patients but charges uninsured patients for the same medicines, we priced their medicines at the price paid by the uninsured.

In the analysis comparing price with quality, we included public sector samples at the price charged to uninsured patients for hospital samples, and at the procurement price for samples from the district warehouse or bought by *puskesmas* with capitation funds.

In comparing maximum retail prices, we kept just one instance of each unique market authorisation number (reflecting a single medicine, dosage, formulation, market authorisation holder and brand or unbranded generic; n=83). In a few cases, because prices may be recalibrated over time, there was more than one maximum retail price per market authorisation number. In these cases, we used the maximum retail price value of the sample with the longest time to expiry, as a proxy for the most recent version of the product.

When calculating the difference between the sales price and the maximum retail price at the sample level, we used the price and maximum retail price for the individual sample.

The analysis code is available at https://doi.org/10.7910/DVN/IQ6HRF.

### Indonesian market data

We extracted data, including market authorisation holder, manufacturer, branded status, brand name (if applicable) for all versions of the study medicines registered for sale in Indonesia from the open-access database maintained by the Indonesian medicine regulator.^27^ With permission from Universitas Pancasila, we merged information on holding companies for market authorisation holders from a database maintained by the Facutly of Pharmacy’s Center for Pharmaceutical Policy and Services Studies, using Stata 17.0 software.

### In-depth interviews

We also conducted in-depth interviews between June 2020 and May 2021 with purposively selected individuals with knowledge of medicine management in public and private sectors (Supplementary Table 1 gives details). Potential participants were approached by e-mail, outlining details of the study and requesting their participation; those who agreed to participate provided verbal or written consent. The interviews, conducted in Indonesian, were recorded and transcribed verbatim. Since the in-depth interviews took place during the COVID-19 pandemic, most interviews were conducted remotely by telephone, WhatsApp, or using the Google Meet platforms.

Topics covered included the influence of price and other incentives of choice of medicines, and perceptions of quality.

### Study Permissions

The Malang District Department of Health gave written permission for the study (070/1102/35.07.103/2020). The study also received ethics approval from the Ethical Committee, Ministry of Research, Technology, and Higher Education, Medical Faculty of Brawijaya University (No.83/EC/KEPK/04/2020) and the Human Research Ethic Committee of University of New South Wales, Sydney (HC200148). Patients were not directly involved in the design, conduct or reporting of this study. The results were reported to the Indonesian medicine regulator within one month of completion of assay and stage 1 dissolution testing.

## Results

In the results section, we integrate data from in-depth interviews where appropriate to provide additional context for quantitative analysis.

### Relationship between price and quality

Figure 1 plots the results of pharmacopeial tests against the relative price of each product to the cheapest for the medicine and dosage. The square markers indicate the medicines provided free to patients in the public health system; these are priced at the public procurement price. All samples met the quality specifications for both assay (Figure 1a) and dissolution (Figure 1b). Thus, there was no relationship between price and quality for any molecule in our study.

**Figure 1 (Colour).**
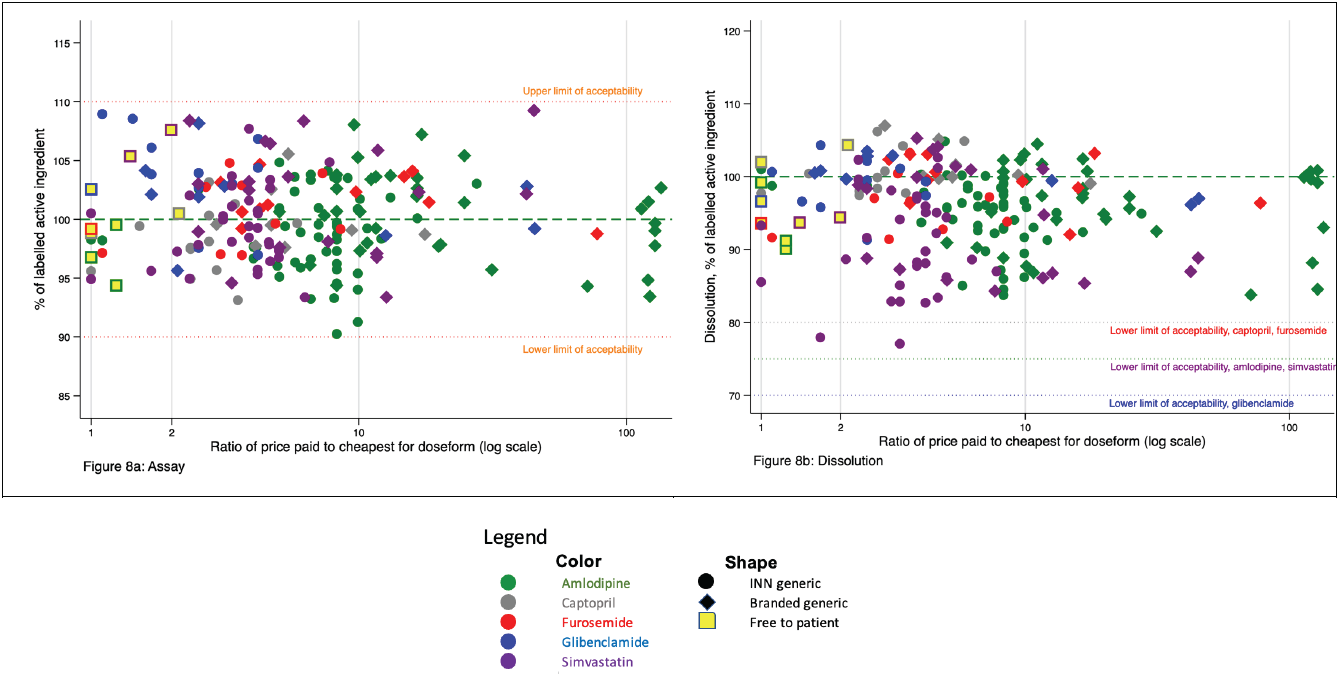
Relationship between price paid and assay (a) and dissolution (b) testing results, of 204 CVD medicines samples, by branded status.

Interviewees were not consistent in their perception of the relationship between quality and price. One doctor, who provided services to poorer patients in the study area shortly before the introduction of the national health insurance programme, was distrustful of low-priced medicines.

*“If I thought a patient was doing ok with a medicine [priced] at 2000 or 3000 per strip [1*.*3-2 cents per pill], then I’d keep them on it. But in several cases, it just didn’t work. So that’s why I had more expensive medicines, branded medicines. So I’d say to the patient “Ma’am, this is a branded medicine, from [Company X] or [Company Y], it costs this much, do you want it or not?” And she’d say “Ya, OK doc, if it’s a good medicine”. And it would work. It worked when she used a branded medicine. So that’s why I don’t know if the contents of that 3000 rupiah medicine is lower than the branded one, or what?”*

Private doctor 1

A public sector pharmacist noted that quality of unbranded products in the public procurement system appeared to have improved in recent years.

*Interviewer: “What’s your opinion of the quality of medicines on e-catalog?”*

*Respondent: It’s variable. I’ve been buying since the beginning, in 2014, and it was all over the place. Like, there was amox[icillin] I think, that each strip only had eight or nine pills, so a box wasn’t 100 [tablets]. it was only 80 or 95*… *There was also antacid that was too thick to pour, and [tablets] that were crushed up, all kinds of stuff. But now, it’s getting better each year, the quality is constantly improving*.

Public Sector Pharmacist.

#### Product variation in the Indonesian market

There were 71 manufacturers making any study medicine at the time of data collection. Some of them produced medicines under contract for several market authorisation holders. A total of 80 market authorisation holders (grouped into 70 holding companies) registered at least one study medicine. Altogether, at the time of the study, there were 110 branded and unbranded versions of amlodipine registered in Indonesia, 56 of simvastatin, 17 of captopril, 13 of furosemide and 12 of glibenclamide.

Because many holding companies sold more than one study medicine, we found a total of 133 holding company-molecule pairs. For example, Holding Company A - amlodipine constitutes one pair, while if Holding Company A also makes simvastatin, this would constitute a second pair.

Looking at holding company - molecule pairs in the Indonesian market, we found that just over half were singletons; either a single brand (35.3%), or a single unbranded (INN) product (15%), see Figure 2. For most of the rest (43.6%), the holding company had registered one branded and one INN version. The remainder registered 2 brands, or 3 or 4 brands/INN versions of the same molecule. In a few cases a single holding company registered a medicine to 2 or 3 different market authorisation holders, allowing them to make more than one INN version of the same medicine.

**Figure 2:**
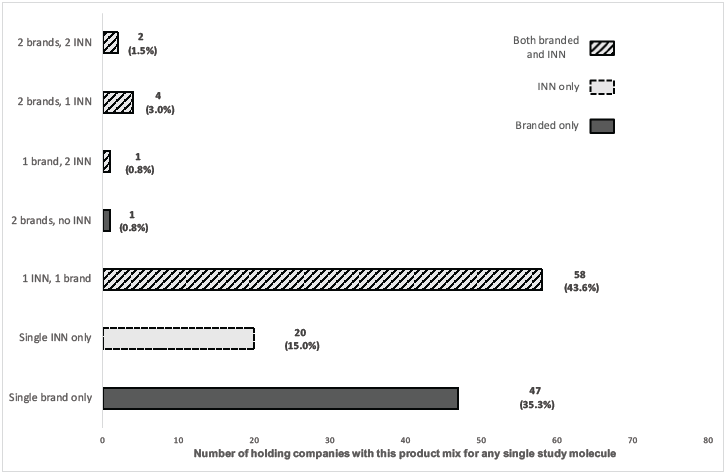
Distribution of product mixes registered, by holding company, for any individual study molecule.

#### Sample distribution in the medicine pricing and quality survey

The sample-level dataset, with brand identities masked in accordance with the terms of our ethical clearance, is available at https://doi.org/10.7910/DVN/IQ6HRF.

Table 2 shows the number of samples and brands by medicine and type of source.

**Table 2:** Details of products collected in study.

|  | Samples |  |  | Brands |  |  | Unique products (brand and dose) |  |  |
| --- | --- | --- | --- | --- | --- | --- | --- | --- | --- |
|  | Branded | INN | Total | Branded | INN | Total | Branded | INN | Total |
| Amlodipine | 35 | 53 | 88 | 14 | 16 | 30 | 18 | 22 | 40 |
| Captopril | 8 | 14 | 22 | 1 | 4 | 5 | 1 | 6 | 7 |
| Furosemide | 11 | 10 | 21 | 4 | 4 | 8 | 4 | 4 | 8 |
| Glibenclamide | 9 | 12 | 21 | 4 | 4 | 8 | 4 | 2 | 6 |
| Simvastatin | 19 | 33 | 52 | 6 | 11 | 17 | 7 | 15 | 22 |
| **Total** | **82** | **122** | **204** | **29** | **39** | **68** | **36** | **49** | **83** |

In accordance with the sampling strategy reflecting population exposure, unbranded generics outnumbered branded generics in both number of samples and variety of products.

#### Variation in maximum retail price of collected samples

Figure 3 shows the value of the maximum retail price for each of the products in the study relative to the lowest maximum retail price for that API and dose. We use a log scale to preserve detail at the lower price ratios. Manufacturers of originator medicines set their highest retail price at between 35 and 75 times that of the cheapest unbranded generic.

**Figure 3:**
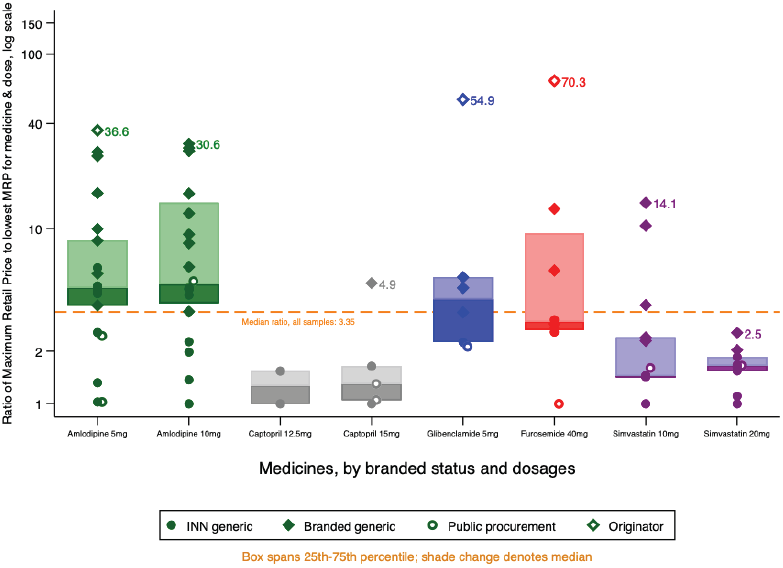
Maximum retail prices, by medicine and branded status.

Maximum retail prices varied considerable even between unbranded (INN) generics. For furosemide and both doses of amlodipine, we found at least one INN generic version with maximum retail prices higher than the lowest maximum retail price for a branded equivalent.

Some pharmacists said in interviews that brand loyalty was strong even for “unbranded” generic products, because patients preferred to stick with manufacturers they knew.

*“Lots of patients are, like, if they’re using [an unbranded generic made by] K[imia] F[arma], they don’t want to use Hexpharm. Or if they’re using Hexpharm, they don’t want to use Dexa. So I have to provide all the versions, I have from Dexa, I have from Hexpharm, I have from Kimia Farma. Though really, ah, those three medicines, the content is the same, the composition is the same, the type is the same, but the price can be quite significantly different*.*”*

Pharmacist 1, private pharmacy

Among the study medicines were samples from 4 holding companies that reflected the range of similar products (same medicine and dosage) they marketed. For 3 of these companies, we sampled both INN and branded generics, while from a fourth we found both an INN generic and 2 branded products.

Figure 4 shows the varied maximum retail prices set by these companies for their similar products. In this case, we show absolute values rather than ratios in order to preserve information about variations in base retail prices set for similar products. While Holding Company 44 priced the products similarly (bottom row on Figure 4, the others priced premium products at between twice and 20 times the price of their unbranded products, with a mean ratio of 8.0. At the time of data collection, the exchange rate was US$1 = IDR 14,370.

**Figure 4:**
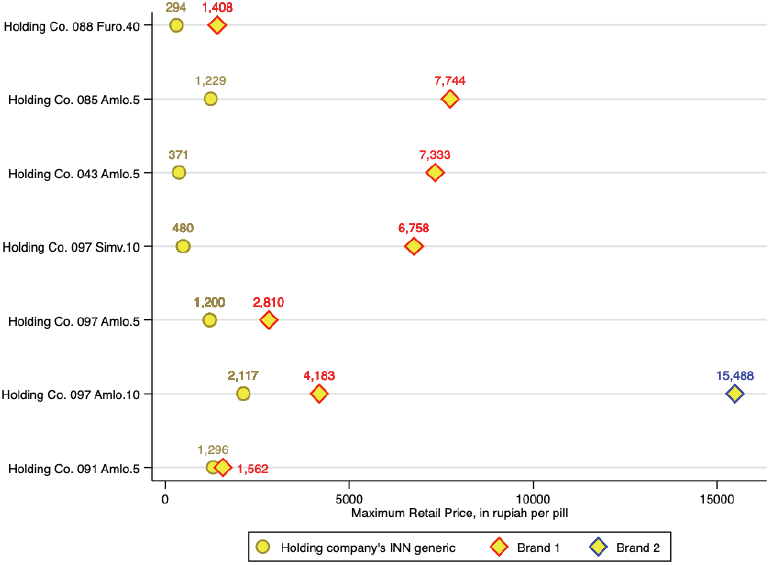
Maximum retail price for branded and unbranded versions of the same product sold by the same company.

#### Variation in retail prices paid

Priced actually paid by patients varied even more than maximum retail prices. Figure 5 shows the ratio of the most expensive to the cheapest price paid for an equivalent product (same API and dose) for all samples that we bought from retail outlets, health providers or the hospital. The graph differentiates by marker shape between unbranded and branded medicines, showing INN generic medicines on the left and their branded equivalent on the right.

**Figure 5:**
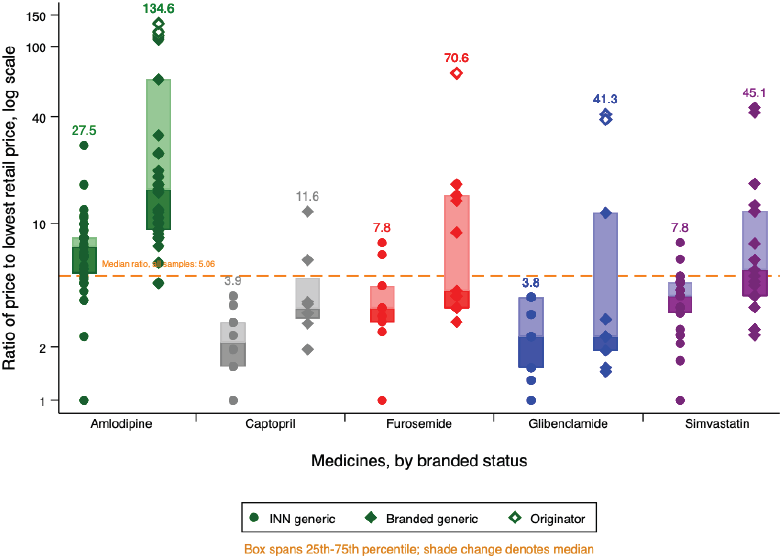
Distribution of relative prices charged, by molecule and branded status.

Five brands of amlodipine (comprising 7 samples) sold at more than 100-fold the price of the cheapest equivalent product, which was an unbranded generic. For all medicines except captopril, there were brands retailing at between 40 and 71 times the cost of the cheapest medicine. As expected from the distribution of maximum retail prices shown in Figure 3, branded medicines were generally more expensive, However, as Figure 5 shows, the relationship was more varied than maximum retail prices would suggest. For all medicines and dosages there were unbranded versions (the top circle marker for each medicine) selling for between 2 and 3.7 times the price of the cheapest branded equivalent (the bottom diamond marker). In all cases, however, the cheapest medicine was INN and the most expensive was branded, with originator brands topping the scale where found. Branded medicines traded in a far wider price range than INN medicines.

Supplementary Table 2 summarises the absolute values underlying the relative values shown in Figure 5, by medicine and dose. Lowest prices for by molecule range from 61 rupiah per tablet for amlodipine 5mg to 239 for simvastatin 20mg (0.4 US cents and 1.7 cents respectively), while the highest prices range from 1,500 rupiah tablet for captopril 15mg to 12,650 for amlodipine 10mg (10.4 cents and 88 cents respectively).

Retailers reported stocking medicine at a variety of price-points, and tailoring their offering to patients based on price signals provided by those patients:

*“Like if a patient asks for [the originator brand], well, that’s expensive, so [if I don’t carry it] I have to find a match that’s more or less the same price*.*”*

Pharmacist 1, private pharmacy

*If they’re asking for an unbranded generic, and we don’t have it, what we have a branded generic but it’s also [priced at] 5,000 [rupiah], they’re fine with that*… *The important thing is that it’s cheap*.

Pharmacist 2, private pharmacy

The price variation was not restricted to the difference between brands. Patients in the Malang area were able to find the identical product (same medicine, dose, formulation and brand or, if unbranded, market authorisation holder) at widely varying prices, as shown in Figure 6.

**Figure 6:**
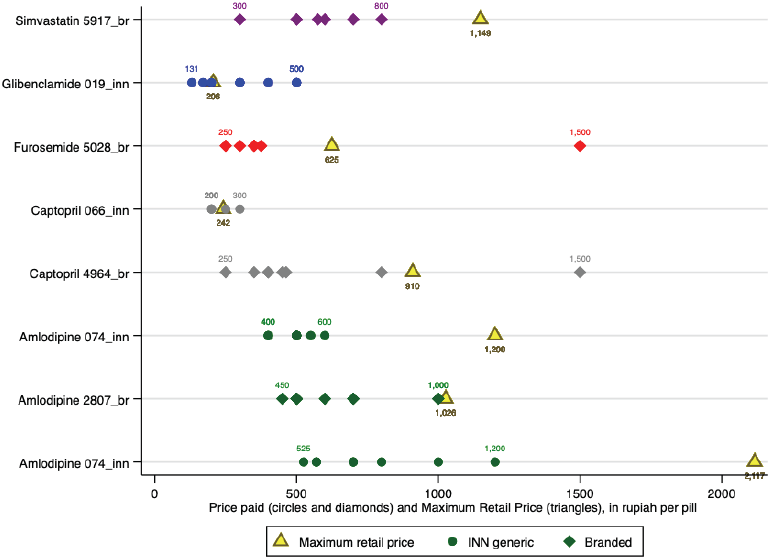
Retail price paid at different outlets, and maximum retail price, for products of the identical medicine, dosage and brand.

Prices for 21% of retail samples exceeded the maximum retail price; this was significantly more common among medicines sold by doctors and midwives, compared with those sold at retail pharmacies/OTC shops or by the hospital (40.7% vs 18.7% and 7.8% respectively, p = 0.016)

Figure 7 shows the distribution of relative prices by medicine and source. Uninsured patients buying unbranded generics from the district hospital benefited from the lowest prices for every medicine. However, hospital patients prescribed or choosing branded medicines paid among the highest prices for those products.

**Figure 7:**
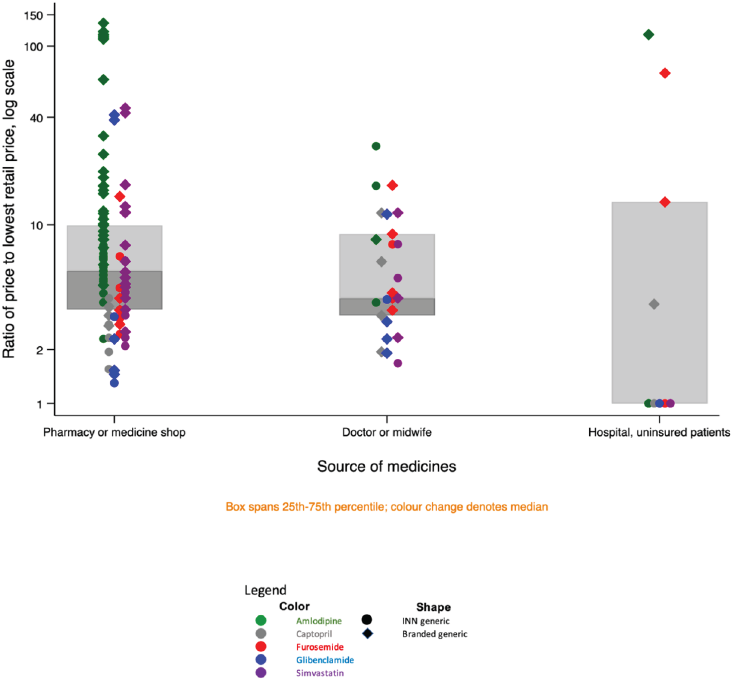
Price variation for similar products, by medicine, branded status and source.

Private doctors and midwives charged a wide range of prices; patients buying from these providers paid among the highest prices for unbranded generics.

### Factors influencing choice of medicines on offer

In interviews, pharmacists and health professionals said that besides wishing to meet patient demand, they decided which products to offer largely on the basis of potential profits or other benefits.

Asked how she chose the distributor or product, one pharmacist replied:

*“Well it depends. Sometimes I chose another [product or distributor], if there’s a something on offer, maybe a bigger discount. If it’s more profitable then I’d choose that*. *\*laughing*”*

Pharmacist 1, private pharmacy.

In public hospitals, pharmacists have the option of buying medicines for insured patients from the national procurement platform, which at the time of the study was a single-winner system with fixed price products. However, they are not obliged to do so. A hospital pharmacist explained that needed products are in any case not always available on the national platform. In that case, hospital pharmacists order according to the hospital formulary, determined based on doctors’ choices.

*“We revise the hospital formulary once every two years, we circulate a list to all the units and doctors: what [brand] do you choose; what do you choose? Then we select three brands, at least the three most common [suggestions]*.*”*

Once a brand is on the hospital formulary, administrators will negotiate for volume-based discounts.

*“The discounts on branded medicines are big, usually, if it’s expensive we can get a big discount So we negotiate, to get the maximum discount. For example, like, Metamizole, it’s supposed to be in the e-catalog but in fact it’s difficult to find, so we negotiate for A*** (a branded version for Metamizole) from the hospital formulary. We negotiate with the producer to give us a maximum discount. At least we try to get the price down close to the e-catalog price, then we can also provide that medicine to the BPJS [publicly-insured] patients*.

Pharmacist 2, public hospital.

Another interviewee, who previously worked as a senior administrator in a private hospital, described intense lobbying of doctors by pharmaceutical companies aiming to get their products on hospital formularies.

*So in private hospitals* … *the deal works like this: the (pharmaceutical company) sales staff will give a percent to the hospital and a percent to the doctor. As I remember the hospital got 15%. Of the price of the medicine [paid by the patient]. The doctor got 20%, 10% [downpayment] up front and 10% based on their monthly prescription values. So if I prescribe 10 million worth of [Company X] products, then I get a million in cash that month. That’s for general practitioners; for specialists, the cut is a lot higher*.*”*

Doctor and former private hospital administrator

Individual health care workers also reported being incentivised to provide specific brands.

*“[Sales staff] will usually give you some kind of household appliance, like an oven. It’s like: Ma’am, if you take these pills, this many boxes, for a few months, then you’ll get this [reward]*.*”*

Private midwife 1

Panel data from 2 private pharmacies confirmed that on average, discounts on branded products were higher than on INN generics. As Table 3 shows, estimated discount averaged 49% for unbranded generics and 62% for branded products. However, because both margins and sale volumes were higher for unbranded generics, these were more profitable for the pharmacies in question.

**Table 3:** Sales data for all versions of study medicines sold at 2 pharmacies, Malang District.

|  | Mean |  |  | Total |  |
| --- | --- | --- | --- | --- | --- |
|  | % discount | Margin (rupiah/pill) | Margin (ratio sell: buy price) | Monthly sales, units* | Monthly profits, rupiah* |
| Branded(n=8) | 62.8 | 82.6 | 1.3 | 737 | 41,878 |
| Unbranded (n=10) | 49.0 | 223.9 | 2.6 | 4,563.4 | 915,769.6 |
| Total | 55.2 | 161.1 | 2.0 | 5,300.4 | 957,647.6 |

\*Average, January-October 2021

## Discussion

Our study recorded a wide variety of prices for similar cardiovascular and anti-diabetic medicines sold to patients in a largely rural district of East Java, but showed that there was no relationship between price and quality. The cheapest medicines all met pharmacopeial specifications, as did equivalent medicines selling at over 100 times the price.

The price differentials in the market suggest that both producers and retailers price products at what they believe the market will bear. They beg the question: why do Indonesian patients pay vastly higher prices for premium medicines when they could get products that meet the same pharmacopeial standards at a fraction of the price (or for free, from public services)?

Interviewees provided some evidence that patient choice is influenced by the suggestions of doctors or other health care providers. Further, they reported that those actors are sometimes incentivised to dispense or prescribe premium brands by the marketing departments of pharmaceutical companies, in contravention of the code of conduct of GP Farmasi, the Indonesian pharmaceutical trade association.^28^ Qualitative research in other parts of Indonesia suggest that doctors sometimes also have a financial interest in local pharmacies, and prescribe to boost profits.^29^

The role of pharmaceutical companies in influencing doctors’ prescribing behaviour is evident even in countries with universal health coverage. In France, the competition authority in 2013 fined Sanofi-Aventis over E 40 million for running a smear campaign against generic versions of clopidogrel, which would compete with its Plavix brand.^30^ In other markets where physicians and hospitals have historically profited from the sale of medicines to patients paying out of pocket, such as the United States and China, physicians continue to express distrust of generic medicines (including for cardiovascular disease) despite large scale studies showing that clinical outcomes do not vary by branding status or price.^4–6,31–34^

In contrast with dynamics in the Indian market reported over a decade ago by Singal and colleagues, ^35^ pharmacists in our study reported receiving bigger discounts from manufacturers of premium products, potentially allowing them to reap significant margins by selling these products. While the hospital reported negotiating large discounts on branded products, high prices paid by uninsured patients for branded study medicines suggest that discounts are not passed on to consumers. According to Kaplan and colleagues, this sort of profit-seeking among those dispensing medicines is common in many settings, and stands in the way of successful implementation of policies designed to promote greater use of cost-effective generic medicines.^36^

In our study, however, several pharmacists reported trying to provide products at the varying price points demanded by different patients. We were unable to interview patients directly -- a major limitation of our study. However, it seems likely that at least some of the demand for premium products in this largely rural area of East Java comes directly from patients because they believe that more money buys better medicine. This would be entirely consistent with the situation reported in other studies, many summarised by Dunne and Dunne.^37^ Poorer patients with limited education are most distrustful of low-cost medicines, and especially of those provided free in the public sector. ^7–9^

The World Health Organization asserted in 2017 that one in 10 medicines in low and middle income countries is substandard or falsified ^39^. However, the few medicine quality surveys that report a relationship between price and quality do not suggest that cheaper or INN generic medicines are any more likely to be poor quality than more expensive or branded medicines. Testing non-communicable disease medicines in Cambodia between 2011 and 2013, Rahman and colleagues found that non-compliant samples of glibenclamide were twice as expensive, on average as those that passed testing, while for amlodipine there was no relationship.^40^ A small study comparing premium (“branded”) and non-premium (“branded-generic”) pairs of medicines produced by the same manufacturer in the Indian market found that all met quality standards.^35^ Among 92 samples of 12 essential medicines collected in Togo, samples that were relatively cheaper compared with an international standard were not significantly more likely to fail than relatively expensive samples.^41^

In our study, one doctor, practicing some years ago, described witnessing treatment failure in patients using low-cost unbranded products, which resolved after switching them to branded products. After the introduction of the national health insurance scheme JKN, and with it free medicines in public facilities, the Indonesian press regularly reported concerns about the quality of those medicines.^17,42^ While in our study a government pharmacist reported a steady reduction in quality problems in recent years, it is possible that perceptions of ineffective or otherwise poor quality cheap medicines rooted in experience have survived, even as actual product quality has improved through investment in production processes and better regulation. Studies in the United States and New Zealand have also demonstrated that the perception of quality equated with price can have a strong placebo effect, even in the absence of active ingredients.^43–45^

Many of the prices in the Indonesian market, including a majority of those in the public procurement system, are now well below the last iteration of the international reference price, adjusted for inflation.^46^ Among 83 unique versions of 4 cardiovascular and one anti-diabetic medicine in Indonesia, comprising a total of 204 samples collected to reflect the likelihood of patient exposure to particular products in a district in East Java, we found no factual basis for any lingering perception that cheap medicines are poor quality. In a large, economically and socially diverse market such as Indonesia’s, it is possible that the quality of low-cost items is protected in part through cross-subsidisation on the part of manufacturers. Regulatory data indicate that it is extremely common for Indonesian companies to register more than one version of the same product -- most commonly a branded and an unbranded version from a single market authorisation holder. Under Indonesian regulations, these must be identical in composition. However, market authorisation holders are free to set their own maximum retail prices for branded products. The wide variation in maximum retail prices suggests companies are engaging in market segmentation, providing products at different price points in order to increase their overall share of the market. In the “paired” products in our study area (INN and branded versions of the same product from the same holding company), companies set prices up to twenty times higher from their branded products compared with the unbranded generic equivalent.

From monthly data recorded by 2 pharmacies, we estimated the discounts at which pharmacies acquire medicines, and found that they were greatest for branded medicines. However, the margin percentage on unbranded generics was on average double that on branded medicines; coupled with higher sales volumes for INN generics, this made these unbranded products more profitable. This observation is based on limited data; we could not verify the wholesale prices at which most pharmacies acquired medicines. However, price variation between pharmacies for the identical product suggests that pharmacists also adapt sales prices to achieve maximum profits, for example by charging lower margins on higher-priced products or those acquired at a greater discount, while pushing up margins on lower-priced, higher volume brands.

In practice, prices charged to consumers were lower than the maximum retail price four-fifths of the time; those charging over the maximum retail price were often unregulated dispensers (doctors, midwives, and medicine shops), who may buy their own medicine supply from retail pharmacies, and who are not subject to oversight by management boards, as is the case in many health facilities and chain pharmacies.

Some politicians, including a former health minister, claim that high-priced medicines are an enduring burden for Indonesian patients.^47^ However, for the 5 study medicines, among the most commonly consumed in Indonesia, we found that for those unwilling or unable to access free medicines from public services, very low cost, quality assured products are widely available in pharmacies. The simultaneous existence of high-priced alternatives for those willing and able to pay for the illusion of better quality is thus not problematic. Indeed, if producers sell expensive branded products to wealthier Indonesians and use some of the profits thus generated to support the sale of very low-cost equivalent INN products to the public health system and to poorer patients, the price variation in the market may benefit health for all Indonesians.

We note that we were only able to test for identity, percent of labelled assay, and dissolution. It is possible that the tested medicines were not uniform in content, contained impurities, or suffered from other defects, and that these defects were more common among lower-priced medicines. In addition, the study was carried out in a largely rural area of the most developed and densely populated island, Java; it is possible that lower-priced medicines degrade more rapidly than those sold at higher prices, and are thus more likely to sink below the quality threshold as they get transported thousands of kilometres to the country’s outer islands.

Overall, however, we conclude that the millions of Indonesian patients who take amlodipine, captopril, furosemide, glibenclamide or simvastatin daily can be broadly confident that their medicines are of acceptable quality, regardless of the price they paid.

## Supporting information

Supplemental Table 1: xlsx format

## Data Availability

Underlying data and analytic code are provided in the Dataverse repository at: https://doi.org/10.7910/DVN/IQ6HRF

## Acknowledgements

We thank Malang District Health Authority, the study participants, and the members of an informal Indonesian medicine supply chain study group whose discussions enriched our understanding of the context. We also thank Slamet Hariono and Elmi Kamilah for their support with data collection. We thank United States Pharmacopeia for providing testing standards at a discounted price for academic research.

## Data availability statement

The sample level data and analysis code are available at https://doi.org/10.7910/DVN/IQ6HRF. In accordance with the terms of the ethics approval, names of individual manufacturers and batch numbers are masked, product registration numbers removed, and outlets grouped by type. Data are provided for reuse, with the expectation that users will cite the dataset and this paper.

