## Supplemental Table 1 for "Variation in price of cardiovascular and diabetes medicine in Indonesia, and relationship with quality: a mixed methods study in East Java"

**Supplementary Table 1: Profile of In-depth Interview contributors**

| Contributor | Sector | Number of individuals interviewed |
|---|---|---|
| **District Warehouse** | Public | 2 |
| **Primary health centre (*Puskesmas*)** | Public | 4 |
| **Community health outpost** | Public | 2 |
| **District hospital** | Public | 1 |
| **Local medicine distributor** | Private | 1 |
| **Independent pharmacy** | Private | 3 |
| **National chain pharmacy** | Private | 2 |
