## Supplemental Table 2 for "Variation in price of cardiovascular and diabetes medicine in Indonesia, and relationship with quality: a mixed methods study in East Java"

**Supplementary Table 2: Details of medicine prices, by medicine and dosage**

| | Prices in Indonesian Rupiah | | | | | Prices in US$ | | | | | |
|---|---|---|---|---|---|---|---|---|---|---|---|
| | Lowest | 25th percentile | Median | 75th percentile | Highest | Lowest | 25th percentile | Median | 75th percentile | Highest | # samples |
| Branded | | | | | | | | | | | |
| Amlodipine 5mg | 450 | 500 | 1000 | 1500 | 8145 | 0.0313 | 0.0348 | 0.0696 | 0.1044 | 0.5668 | 19 |
| Amlodipine 10mg | 500 | 1000 | 1664 | 12000 | 12650 | 0.0348 | 0.0696 | 0.1158 | 0.8351 | 0.8803 | 16 |
| Captopril 15mg | 250 | 375 | 425 | 632 | 1500 | 0.0174 | 0.0261 | 0.0296 | 0.0439 | 0.1044 | 8 |
| Glibenclamide 5m | 190 | 250 | 300 | 1500 | 5400 | 0.0132 | 0.0174 | 0.0209 | 0.1044 | 0.3758 | 9 |
| Furosemide 40mg | 250 | 300 | 375 | 1300 | 6369 | 0.0174 | 0.0209 | 0.0261 | 0.0905 | 0.4432 | 11 |
| Simvastatin 10mg | 300 | 575 | 750 | 1513 | 5800 | 0.0209 | 0.0400 | 0.0522 | 0.1053 | 0.4036 | 14 |
| Simvastatin 20mg | 600 | 600 | 800 | 1833 | 4000 | 0.0418 | 0.0418 | 0.0557 | 0.1276 | 0.2784 | 5 |
| Unbranded | | | | | | | | | | | |
| Amlodipine 5mg | 61 | 400 | 500 | 550 | 1667 | 0.0042 | 0.0278 | 0.0348 | 0.0383 | 0.1160 | 29 |
| Amlodipine 10mg | 109 | 500 | 600 | 750 | 1300 | 0.0076 | 0.0348 | 0.0418 | 0.0522 | 0.0905 | 21 |
| Captopril 12.5mg | 73 | 200 | 200 | 200 | 250 | 0.0051 | 0.0139 | 0.0139 | 0.0139 | 0.0174 | 5 |
| Captopril 15mg | 129 | 200 | 200 | 300 | 500 | 0.0090 | 0.0139 | 0.0139 | 0.0209 | 0.0348 | 7 |
| Glibenclamide 5m | 131 | 200 | 300 | 500 | 500 | 0.0091 | 0.0139 | 0.0209 | 0.0348 | 0.0348 | 11 |
| Furosemide 40mg | 90 | 250 | 300 | 400 | 700 | 0.0063 | 0.0174 | 0.0209 | 0.0278 | 0.0487 | 9 |
| Simvastatin 10mg | 129 | 400 | 500 | 600 | 1000 | 0.0089 | 0.0278 | 0.0348 | 0.0418 | 0.0696 | 14 |
| Simvastatin 20mg | 239 | 800 | 1000 | 1003 | 1500 | 0.0166 | 0.0557 | 0.0696 | 0.0698 | 0.1044 | 17 |
